# Genetically-Determined Low-density Lipoprotein in Ischemic Stroke Survivors

**DOI:** 10.1101/2025.02.26.25322908

**Authors:** Kane Wu, Shufan Huo, Cyprien A. Rivier, Victor M. Torres-Lopez, Richa Sharma, Adam de Havenon, Bradford Worrall, Kevin N. Sheth, Guido J. Falcone

**Affiliations:** Department of Neurology, Yale School of Medicine, New Haven, CT, USA; Department of Neurology, University of Virginia.

**Author notes:** **Corresponding Author:** Guido J. Falcone, MD ScD MPH, 15 York Street, LLCI Room 1004D, P.O. Box 208018, New Haven, CT 06510, USA. These authors contributed equally (Kane Wu, Shufan Huo).

## Abstract

**Background and Objectives:** Polygenic susceptibility to hypertension and diabetes negatively impacts the clinical trajectory of ischemic stroke survivors. We hypothesize that polygenic susceptibility to hyperlipidemia (PSH) negatively impacts cholesterol control in this same population.

**Methods:** We conducted a genetic association study using data from the Vitamin Intervention Stroke Prevention (VISP) study, a clinical trial that enrolled survivors of ischemic stroke. PSH was modeled through a polygenic risk score built with 38 independent genetic risk variants for LDL-c that was divided into <20, 20-80, and >80 percentile categories labeled as low, intermediate and high PSH. We used multivariable linear, logistic and Cox regression, as appropriate, to test whether high PSH was associated with risk of uncontrolled hyperlipidemia (LDL-c > 100mg/dl), resistant hyperlipidemia (LDL-c > 100mg/dl despite statin treatment) and clinical outcomes. We replicated our findings in a cohort of ischemic stroke survivors enrolled in the UK Biobank.

**Results:** 1,567 ischemic stroke survivors (mean age 68 years, 35% female) enrolled in VISP were included in the study. Stroke survivors with higher versus low PSH had 66% higher risk of uncontrolled hyperlipidemia (OR 1.66, 95%CI 1.17-2.36), 83% higher risk of resistant hyperlipidemia (1.83, 95%CI 1.00-3.37), twice the risk of recurrent stroke (HR 2.03, 95%CI 1.14-3.61) and 80% higher risk of acute coronary events (HR 1.81, 95%CI 1.18-2.78). The association between high PSH and higher risk of uncontrolled and resistant hyperlipidemia were replicated in 1,634 stroke survivors (mean age 61, 32% female sex) enrolled in the UK Biobank (OR 2.34, 95%CI 1.68-3.28 and OR 2.33, 95%CI 1.61-3.38, respectively).

**Discussion:** Among acute ischemic stroke survivors, higher PSH is associated with worse lipid control and higher risk of recurrent vascular events. Combined with existing evidence on the role of adverse genetic profiles in blood pressure and blood control in this population, our findings support the comprehensive evaluation of polygenic profiles as a cause of failed risk factor control in stroke survivors.

## INTRODUCTION

Stroke is one of the top five leading causes of death and disability in the United States and worldwide.^1^ With advancements in treatment and rehabilitation, the number of stroke survivors has steadily increased.^2^ This vulnerable group is at particularly high risk of stroke recurrence, cognitive decline, and dementia.^3,4^ These secondary events cause a high additional burden of disability, morbidity, and mortality for more than 70% of stroke survivors within five years.^5^

Elevated low-density lipoprotein cholesterol (LDL-c) is one of the main treatable risk factors for both first-time stroke and stroke recurrence, acute coronary events and dementia in stroke survivors.^6,7^ While lipid lowering treatments are generally shown to reduce mortality and morbidity, efficacy and outcomes vary due to polygenetic variations and environmental factors influencing LDL-c elevation.^8,9^ Current guidelines for antihyperlipidemic therapy considering classical cardiovascular risk factors such as age, blood pressure, and cholesterol measurements often fail to account for these inter-individual differences.^10^

Genetic variation explains around 30-40% of the variance of LDL-c levels in the general population.^11^ This genetic contribution towards elevated LDL-c is a synergy of many common genetic risk variants. The aggregate burden of these variants carried by an individual determines their overall polygenic susceptibility to hyperlipidemia (PSH). Prior research indicates that high PSH is associated with higher risk of first-ever cardiovascular events;^12^ however, research on the role of PSH after stroke remains limited. Describing genetic risk factors for post-stroke hyperlipidemia and secondary events can enable the development of individually tailored prevention strategies in stroke aftercare. We hypothesize that genetically elevated LDL-c levels negatively impact antihyperlipidemic treatment control and clinical trajectories after stroke, leading to higher risk of recurrent stroke, coronary events, and death.

## METHODS

### Study design and participant selection

We conducted a 2-stage (testing and replication) genetic association study. For the primary stage, we conducted a post-hoc analysis of clinical and genetic data collected for the Vitamin Intervention Stroke Prevention (VISP) trial, which examined high-dose vitamins in ischemic stroke survivors. VISP is a multi-center, double-blind, randomized, controlled clinical trial enrolling patients aged 35 or older with a non-disabling ischemic stroke within 120 days and homocysteine levels above the 25th percentile at screening. Ischemic stroke was defined as a non-cardioembolic ischemic stroke with a neurological deficit persisting for at least 24 hours, or an infarction in the corresponding brain region confirmed by neuroimaging. Non-disabling was defined as a modified Rankin score ≤ 3. Participants were randomized to receive either a low or high dose daily multivitamin containing pyridoxine (Vitamin B6), folic acid (Vitamin B9), and cobalamin (Vitamin B12) and were followed for a two-year period at three-month interval clinic and phone appointments to assess interim recurrent stroke. Incidences of other vascular events (myocardial Infarction, coronary artery bypass grafting, etc.) were additionally tracked by physician assessment, hospital records, and reviewed by committee. The protocols for the VISP trial are further described elsewhere.^13^ We evaluated data from the nested genetic cohort study “Genomics and Randomized Trials Network” (GARNET) completed in 2,164 participants of the VISP trial. Because the majority of enrolled participants were of European ancestry, we further limited genetic analysis to participants of European ancestry to minimize population stratification effects and genetic confounding.

### Genomic data

Genome-wide data were generated using the Illumina’s HumanOmni1-Quad array and genotyping was conducted at the Center for Inherited Disease Research, Johns Hopkins University. We performed standard quality control procedures for genome-wide data including genotype level filters testing for departure from Hardy-Weinberg equilibrium, genotype call rates, sex effects, and missingness between cases.^14^ Subject level quality checks included relatedness and heterozygosity. Population stratification was conducted to assess for European genetic ancestry.^15^ This pipeline yielded 1,761 subjects and 791,723 markers that were imputed to 1000 Genomes integrated reference panels (phase 3 integrated variant set release in NCBI build 37)^16^ using SHAPEIT^17^ and IMPUTE2^18^ after quality control and multidimensional scaling.

### Exposure of interest

Our exposure of interest was PSH modeled through a polygenic risk score for LDL-c levels. Polygenic risk scores (PRS) constitute a well-established tool in population genetics that estimates an individual’s genetic burden across numerous genetic risk variants.^19^ For a given study participant, the PRS is the sum of the product of the risk allele counts for each variant multiplied by the allele’s reported effect on LDL-c levels. We built a PRS using genetic information on 38 known genetic risk variants, also called single nucleotide polymorphisms (SNPs), associated with higher LDL-c. To isolate LDL-c specific effects, these genetic variants were only associated with higher LDL-c levels and not with other plasma lipid components or other traits.^11^ In addition, these SNPs had a minor allele frequency >1% and were independent (r2 <0.1), biallelic (involve two alleles only) and associated with increased plasma LDL-c at genome-wide levels (p<5×10^-8^). To represent the PSH as a categorical variable in an intuitive manner, we divided participants into <20, 20-80, and >80 percentile categories labeled as low, intermediate and high PSH.

### Outcomes of interest

We evaluated several primary outcomes representing lipid control in stroke survivors: plasma LDL-c at enrollment, uncontrolled hyperlipidemia, and treatment-resistant hyperlipidemia. We defined uncontrolled hyperlipidemia as baseline LDL > 100 mg/dL at enrollment and resistant hyperlipidemia as LDL > 100 mg/dL despite being on statin treatment according to current therapeutic guidelines.^20–22^ The detailed description of LDL cholesterol measurements in the VISP trial has been published previously.^13,23^ Briefly, plasma LDL-c was obtained during the baseline examination. To determine whether patients were on statin treatment, we used data from self-reported information in the baseline interview completed by all participants. Secondary clinical endpoints were stroke recurrence, a composite of stroke recurrence or acute coronary event, and composite of stroke recurrence, acute coronary event, or all-cause death. Qualifying coronary events included hospitalized myocardial infarction, coronary collapse, and coronary artery bypass graft or coronary angioplasty performed. Clinical endpoints and patient deaths were recorded with available information from hospital records, death certificates, coroner’s reports, and physician questionnaires. All clinical outcomes were centrally adjudicated by a panel of clinical experts.

### Covariates

Age at baseline was calculated as the difference between the participant’s birthdate and the date of randomization in the VISP trial and treated as a continuous variable. Sex and ancestry were determined using genetic information. Statin usage, hypertension diagnosis, and diabetes diagnosis as binary categorical variables were identified by combining the baseline forms and physical exam data.

### Statistical analysis

#### Primary analysis

First, we investigated unadjusted associations between PSH and LDL-c levels, uncontrolled hyperlipidemia and treatment-resistant hyperlipidemia using unpaired t, ANOVA, and chi-square tests as appropriate. Second, to account for bias introduced by potential confounders, we also evaluated these same relationships using multivariable linear (LDL-c levels) and logistic regression (uncontrolled and treatment-resistant hyperlipidemia) models that were adjusted for age, sex, hypertension, diabetes, and statin treatment. All variables were evaluated to identify data entry errors and participants with missing exposures or outcomes of interest were excluded from analysis.

#### Replication

To maximize the scientific rigor of this study, we replicated our primary analyses using data from the UK Biobank, a population-based cohort study that enrolled around 500,000 participants throughout the United Kingdom.^24^ Sociodemographic information, questionnaires, physical measurements, information about medication intake including statins, and bio-samples including blood samples for LDL-c measurements and genotyping were collected at baseline. Clinical data including incident ischemic stroke before and after baseline were retrieved via electronic health records and ICD-9/10 codes. We included participants of European ancestry with ischemic stroke prior to enrollment. The statistical analyses for correlation between PSH and LDL-c, uncontrolled and resistant hyperlipidemia were performed equivalent to the discovery cohort.

#### Secondary analysis

We used Cox proportional hazards models and survival analyses via the log rank test to compare the relative risk of recurrent stroke between the intermediate and high PSH categories versus low PSH, repeating the analysis for composite endpoints of ischemic stroke and coronary event, as well as a composite including all-cause mortality. Participants were censored at loss-to-follow-up and the date was set from the VISP trial data. We confirmed the proportional hazard assumption over time by calculating Schoenfeld residuals for all variables in our multivariable models. We adjusted for age, sex, hypertension, diabetes, and statin treatment in multivariable Cox proportional hazards models to test whether higher PSH was associated with stroke recurrence and the composite endpoints. As a sensitivity analysis, we calculated the cox proportional hazards for clinical endpoints while including measured LDL as a covariable. This sensitivity adjusted model would investigate whether the PSH increased the risk of recurrent stroke beyond the risk captured by measured LDL at the time of enrollment.

#### Statistical significance and software

We declared statistical significance at p<0.05. We used PLINK (version 1.9) to conduct quality control procedures and polygenic risk scoring, and R (version 3.6) for all other analyses.

### Standard Protocol Approvals, Registrations, and Patient Consents

The VISP trial protocols were approved by the ethics committees of all study institutions and administrative sites with written informed consent obtained from every potential participant prior to screening. Ethics approval for the UK Biobank study was obtained from the North West Centre for Research Ethics Committee and all participants or their legal guardians have provided written informed consent.

### Data availability

The data from GARNET are publicly available on the database of Genotypes and Phenotypes (dbGaP) and was acquired through the accession number phs000343.v3.p1. Data from the UK Biobank are publicly available and was accessed using project application number 58743.

## RESULTS

Of the total 3680 participants enrolled in VISP, 2,164 participants were enrolled in GARNET and had available genetic data. Of these, 19 (0.9%) were excluded during the genomic quality control process and 342 (16.3%) participants were excluded for non-European ancestry. Of the resulting 1761 participants, one was rejected after imputation and ten rejected for ID mismatch (0.6%). Out of 1750 remaining participants, 1567 (89.5%) had a recorded LDL-c baseline measurement on file and were thus included in this study (mean age 68 years [SD 10.7]; 549 (35%) were female, Figure 1). The mean LDL-c in this final analytical cohort was 122.6 mg/dL [SD 38] (Table 1). The proportion of uncontrolled and resistant hyperlipidemia in the cohort was 71% (n=1110) and 23% (n=355), respectively (Supplemental Table 2).

**Figure 1.**
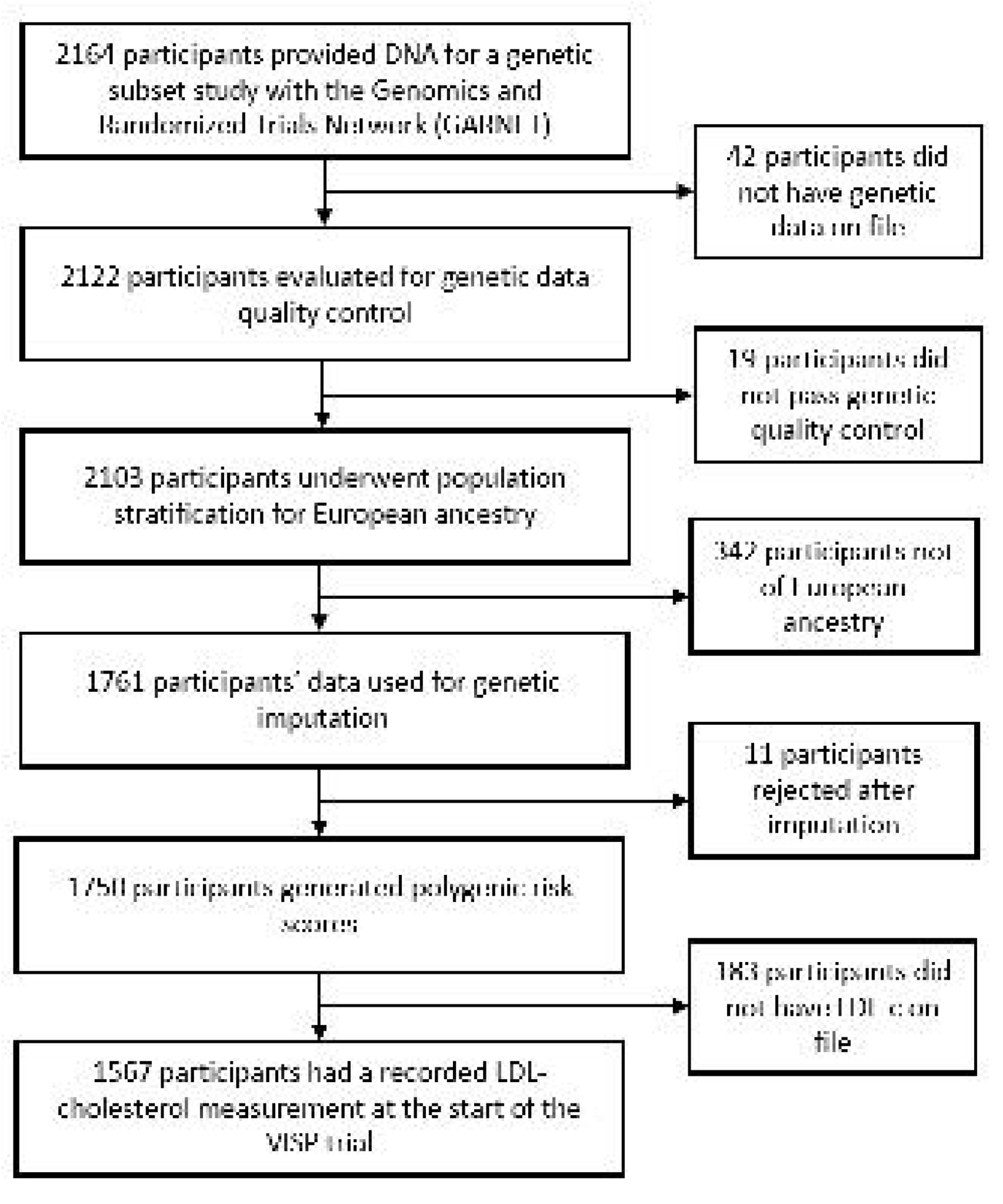
Cohort Selection

**Table 1.** Baseline Characteristics from selected VISP cohort.

| Variable | Polygenic susceptibility to high LDL-c |  |  |  |  |
| --- | --- | --- | --- | --- | --- |
|  | All | Low (0-20%) | Intermediate (20-80%) | High (80-100%) | P-value |
|  | n= 1567 | n = 313 | n = 940 | n = 314 |  |
| Variables of interest: |  |  |  |  |  |
| LDL (mean, SD) | 122.64 (38.0) | 114.5 (38.8) | 123.2 (37.5) | 128.8 (41.9) | <0.001 |
| Recurrent stroke (n, %) | 127 (8.1) | 18 (5.7) | 76 (8.1) | 33 (10.5) | 0.088 |
| Demographics: |  |  |  |  |  |
| Age at enrollment (mean, SD) | 67.95 (10.74) | 68.33 (10.53) | 67.84 (10.83) | 67.9 (10.70) | 0.613 |
| Female sex (n, %) | 549 (35.0) | 103 (32.8) | 342 (36.4) | 104 (33.2) | 0.389 |
| Vascular risk factors: |  |  |  |  |  |
| Hypertension (n, %) | 1089 (69.4) | 220 (70.1) | 659 (70.3) | 210 (67.3) | 0.607 |
| Diabetes Mellitus (n, %) | 385 (24.6) | 87 (27.7) | 221 (23.5) | 77 (24.6) | 0.327 |
| Smoking (n, %) | 221 (14.1) | 43 (13.7) | 138 (14.7) | 40 (12.8) | 0.686 |
| BMI (mean, SD) | 28.0 (5.5) | 27.8 (4.8) | 28.1 (5.8) | 27.7 (5.1) | 0.818 |
| Comorbidities: |  |  |  |  |  |
| Prior MI (n, %) | 277 (17.7) | 53 (16.9) | 164 (17.4) | 60 (19.2) | 0.722 |
| CHF (n, %) | 78 (5.0) | 13 (4.1) | 49 (5.2) | 16 (5.1) | 0.753 |
| Cancer (n, %) | 212 (13.5) | 48 (15.3) | 120 (12.8) | 44 (14.1) | 0.496 |
| Prior CABG (n, %) | 168 (10.7) | 27 (8.6) | 104 (11.1) | 37 (11.8) | 0.383 |
| Medications: |  |  |  |  |  |
| Statin use (n, %) | 517 (33.0) | 94 (30.0) | 314 (33.4) | 109 (34.7) | 0.407 |
| Antiplatelet therapy (n, %) | 397 (25.3) | 87 (27.7) | 238 (25.3) | 72 (23.0) | 0.400 |

Abbreviations: SD = Standard Deviation. BMI = Body Mass Index. MI = Myocardial Infarction. CHF = Congestive Heart Failure. CABG = Coronary Artery Bypass Graft

### Polygenic susceptibility to hyperlipidemia and observed LDL

Higher PSH, modeled via PRS, was significantly associated with LDL-c levels in stroke survivors. When comparing study participants with low, intermediate, and high PSH, the mean LDL-c was 114.5 [SD 38.8], 123.2 [SD 37.5] and 128.8 [SD 41.9], respectively (unadjusted p<0.001) (Table 1). These associations remained significant in multivariable linear regression analyses comparing intermediate vs low PSH (beta 7.96, SE 2.51) and high versus low PSH (beta 14.29, SE 3.08; test for trend p < 0.001) (Table 2).

**Table 2.** Polygenic Susceptibility to Hyperlipidemia and LDL-c control.

| Outcome | Analytical Strategy | Genetic Risk Cohort | Garnet-VISP N=1567 |  | UKB N=1634 |  |
| --- | --- | --- | --- | --- | --- | --- |
|  |  |  | Beta (SE) or OR (95% CI) | P trend | Beta (SE) or OR (95% CI) | P trend |
| Measured LDL | Linear Regression | Low | Reference | <0.001 | Reference | <0.001 |
|  |  | Intermediate | 7.96 (2.51) |  | 12.01 (1.81) |  |
|  |  | High | 14.29 (3.08) |  | 15.55 (2.23) |  |
| Uncontrolled Hyperlipidemia | Logistic Regression | Low | Reference | 0.003 | Reference | <0.001 |
|  |  | Intermediate | 1.50 (1.14-1.97) |  | 1.98 (1.50-2.60) |  |
|  |  | High | 1.66 (1.17-2.34) |  | 2.34 (1.68-3.28) |  |
| Resistant Hyperlipidemia | Logistic Regression | Low | Reference | 0.003 | Reference | <0.001 |
|  |  | Intermediate | 1.79 (1.11-2.90) |  | 2.07 (1.52-2.84) |  |
|  |  | High | 1.83 (1.01-3.33) |  | 2.33 (1.61-3.38) |  |

Abbreviations: CI = Confidence Interval. OR = Odds Ratio. SE = Standard Error.

### Polygenic susceptibility to hyperlipidemia and uncontrolled/resistant hyperlipidemia

A higher PSH was also associated with a higher likelihood of uncontrolled hyperlipidemia in stroke survivors. When comparing study participants with low versus high PSH, the proportion with uncontrolled hyperlipidemia was 63.1% (n=198) versus 74.1% (n=232), respectively (p = 0.003). We observed a similar relationship between PSH categories and resistant hyperlipidemia: In patients with low PSH, 17.3% (n=54) had resistant hyperlipidemia, whereas it was 24.2% (n=76) in patients with high PSH (p = 0.03) (Supplemental Table 2). These associations remained significant in the multivariable modeling even after adjusting for covariates (Table 2). Those in the intermediate risk cohort (OR 1.50, 95% CI 1.14-1.97) and high-risk cohort (OR 1.66, 95% CI 1.17-2.34) were more likely to have uncontrolled hyperlipidemia, and the same was true for the likelihood of resistant hyperlipidemia in the intermediate (OR 1.79, 95% CI 1.11-2.90) and high-risk cohorts (OR 1.83, 95% CI 1.01-3.33).

### Replication

Of 502,369 participants enrolled in the UK Biobank, 1,913 had a history of prior ischemic stroke. Of those, 1,838 had genomic data that passed quality control. 1,731 of these participants were white and 1,634 of these had LDL-c measurements and were included in the final analysis (mean age 61 years [SD 6.7], 31.7% females) (Supplemental Table 1). The mean LDL-c was 105.9mg/dl [SD 6.7]. The proportion of uncontrolled and resistant hyperlipidemia in the cohort was 52% (n=847) and 33% (n=537), respectively (Supplemental Table 2). When comparing participants with low, intermediate, and high PSH, the mean LDL-c was 97.2mg/dl [SD 28.4], 107.3mg/dl [SD 31.5] and 110.2mg/dl [SD 33.1], respectively (unadjusted p<0.001). Participants with intermediate (OR 1.98, 95% CI 1.50-2.60) and high genetic risk (OR 2.34, 95% CI 1.68-3.28) were twice as likely to have uncontrolled hyperlipidemia, and the same relationship was observed for resistant hyperlipidemia in the intermediate (OR 2.07, 95% CI 1.52-2.84) and high-risk cohorts (OR 2.33, 95% CI 1.61-3.38). Thus, all results from the primary analysis could be replicated (Table 2).

### Polygenic susceptibility to hyperlipidemia and clinical endpoints

Over a total follow-up period of 2,866 person-years (1.8 years mean follow-up, SD 0.36 years), 127 recurrent stroke endpoints occurred. Compared to stroke survivors with low PSH, the unadjusted cox-proportional hazards model showed that those with intermediate (HR 1.43, 95% CI 0.85-2.39) and high (HR 1.97, 95% CI 1.11-3.48) PSH had a higher risk of stroke recurrence (p-trend 0.02). Similar associations of higher PSH and stroke recurrence were observed in adjusted multivariable hazard modeling for intermediate (HR 1.52, 95% CI 0.91-2.54) and high (HR 2.03, 95% CI 1.14-3.61) risk categories. Differences for the composite endpoint of stroke and acute coronary events were observed for intermediate (HR 1.31, 95% CI 0.90-1.92) and high (HR 1.81, 95% CI 1.18-2.78) risk categories as well (Table 3). Inclusion of all-cause mortality further confirmed the increased hazard of adverse outcomes for intermediate and high PSH groups. These associations persisted when examining survival curves with the log rank test in adjusted analyses for recurrent stroke and composite endpoints (p<0.001) (Figure 2). Sensitivity analysis showed similar increases in risk of stroke recurrence in intermediate (HR 1.51, 95% CI 0.90-2.53) and high PSH groups (HR 2.00, 95% CI 1.12-3.57) even when including measured baseline LDL in the proportional hazard model, which was confirmed for both composite endpoints (Table 3). Schoenfeld residuals showed that cox proportional hazard modeling assumptions were met in all analyses.

**Figure 2.**
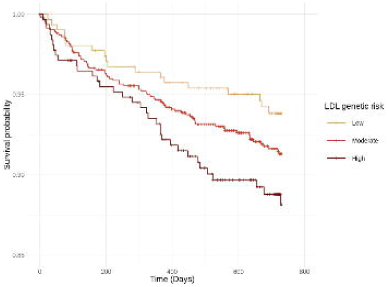
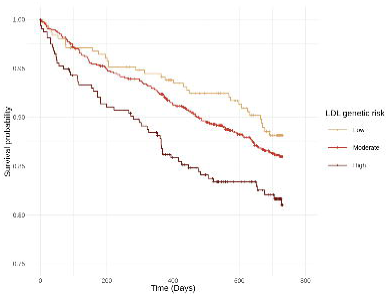
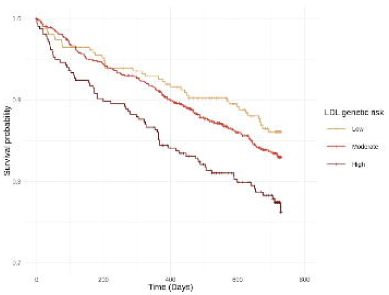
Survival Curves (A) Recurrent Stroke by Polygenic Susceptibility to Hyperlipidemia. (B) Recurrent Stroke or Coronary Event by Polygenic Susceptibility to Hyperlipidemia. (C) Recurrent Stroke or Coronary Event or Death by Polygenic Susceptibility to Hyperlipidemia.

**Table 3.** Clinical Endpoints in VISP cohort.

| Genetic Risk Cohort | Unadjusted HR(CI) | P trend | adjusted HR(CI) | P trend | Sensitivity adj. HR(CI) | P trend |
| --- | --- | --- | --- | --- | --- | --- |
| Recurrent Stroke |  |  |  |  |  |  |
| Low | Reference | 0.02 | Reference | 0.01 | Reference | 0.02 |
| Intermediate | 1.43 (0.85-2.39) |  | 1.52 (0.91-2.54) |  | 1.51 (0.90-2.53) |  |
| High | 1.97 (1.11-3.48) * |  | 2.03 (1.14-3.61) * |  | 2.00 (1.12-3.57) * |  |
| Recurrent Stroke or Coronary Event |  |  |  |  |  |  |
| Low | Reference | <0.01 | Reference | <0.01 | Reference | 0.01 |
| Intermediate | 1.22 (0.83-1.79) |  | 1.31 (0.90-1.92) |  | 1.28 (0.88-1.88) |  |
| High | 1.75 (1.14-2.68) * |  | 1.81 (1.18-2.78) * |  | 1.72 (1.12-2.65) * |  |
| Recurrent Stroke, Coronary Event, or Death |  |  |  |  |  |  |
| Low | Reference | <0.01 | Reference | <0.01 | Reference | <0.01 |
| Intermediate | 1.24 (0.88-1.75) |  | 1.33 (0.94-1.88) |  | 1.30 (0.92-1.84) |  |
| High | 1.81 (1.23-2.67) * |  | 1.88 (1.28-2.77) * |  | 1.80 (1.22-2.66) * |  |

Abbreviations: CI = Confidence Interval. HR = Hazard Ratio.

## DISCUSSION

In our study, higher PSH, modeled through PRS, negatively impacted post-stroke LDL-c control, including higher LDL-c values and a greater incidence of uncontrolled and treatment resistant hyperlipidemia. These associations were consistent after adjusting for confounding and when replicated in an independent cohort. Higher PSH also was associated with greater stroke recurrence, cardiovascular morbidity and death. These results were confirmed in our sensitivity analyses, which demonstrated that the predictive value of PSH persisted even when including observed LDL-c measurements of individuals at the time of enrollment in the VISP trial.

Clinical care of stroke survivors can be challenging due to the elevated risk of stroke recurrence, acute coronary events, cognitive decline and dementia.^4,25^ Along with the possibility of stroke-related disabilities, these complications carry substantial additional morbidity and reduction in quality of life. A meta-analysis of randomized control trials concluded that every 1 mmol/L (39 mg/dL) of LDL-c reduction translates into a 20-25% decrease in risk of major cardiovascular events, observed across multiple types of lipid-lowering therapy.^26^ As is the case in conventional guidelines regarding primary prevention, vigilant control of cardiometabolic risk factors in stroke survivors constitute an effective way to reduce these morbidity and mortality burdens.^27^ Despite compelling evidence showing the beneficial effects of lipid optimization strategies, especially when combined with intensive management with other vascular risk factors, an important proportion of stroke survivors have LDL-c levels that are not at goal. A better understanding of the biology and risk factors for elevated LDL-c resistant to statin treatment may lead to better strategies to identify and address stroke survivors at high risk of dangerous LDL-c trajectories.

While the role of PSH in primary occurrence of cerebrovascular diseases has been studied, little is known about its influence on clinical trajectories after stroke. Our findings address this gap in knowledge, not only by highlighting the impact of PSH on LDL-c in stroke patients, but also on the more prominent role of PSH in the subgroups of uncontrolled and resistant hyperlipidemia. Furthermore, we demonstrate that PSH has the capacity to be a greater predictive tool for stroke recurrence than measured LDL-c alone, as PSH represents a person’s lifetime exposure to greater hyperlipidemia risk before and after any single LDL-c measurement from blood. These results, highlighting the adverse influence of PSH on vascular long-term outcome measures, emphasize the need to incorporate genetic factors in risk assessment and treatment decisions after stroke.

While treatment guidelines for other cardiovascular risk factors such as hypertension and diabetes have numerical cutoffs, prescription of statins and other lipid-lowering therapies are dependent on cardiovascular risk assessments^28–30^. These considerations, however, cannot account for inter-individual differences in risk and treatment response.^31^ Integrating genomic data into novel tools could identify stroke survivors who stand to benefit most from earlier and more aggressive treatment for hyperlipidemia. Considering genetic factors is one important component of today’s precision medicine advances. Ultimately, we are moving towards integration of existing approaches for risk assessment using clinical, socioeconomic, and cultural data. Supporting these ideas, genetic studies of the *Further Cardiovascular Outcomes Research With PCSK9 Inhibition in Subjects With Elevated Risk (FOURIER)* and *Evaluation of Cardiovascular Outcomes After an Acute Coronary Syndrome During Treatment With Alirocumab (ODYSSEY OUTCOMES)* trials demonstrated that patients with higher genetic risk of cardiovascular disease have the greatest benefit from aggressive lipid lowering with proprotein convertase subtilisin/kexin type 9 (PCSK-9) inhibitors.^32,33^

Furthermore, the application of both genetic risk and clinical predictive scores could produce better health outcomes than either approaches alone.^34^ Communicating genetic risk in counseling lifestyle interventions addressing cardiovascular diseases could potentially lead to more effective patient education and adherence.^35^ As the financial costs of genotyping decrease and the number of persons with available genome-wide data increase, these strategies hold promising potential as an integrative and widely applicable tool in clinical practice.

Our study has several limitations. Firstly, therapeutic options for hyperlipidemia have increased since the VISP trial was initially designed and conducted. Our analysis only accounts for the modulation of PSH by statin treatment. Furthermore, our data did not differentiate between types of statins, which could obscure differential susceptibility to genetic resistance. Secondly, medication data in the VISP trial and the UK Biobank are self-reported, and medication doses and adherence could not be assessed, which may inflate the estimate of resistant hyperlipidemia in our cohort. Clinical outcomes analysis was not replicated due to limited data for secondary events. Finally, we could only include participants from European genetic ancestry, which limits the generalizability of our results to other ancestry groups. The importance of external validation in persons from different ancestries cannot be understated, as the underrepresentation of minority populations in genetic studies is widely known. Uncritical use of this data for precision medicine tools could worsen existing health disparities and should therefore be avoided.

In summary, PSH is associated with worse lipid control and clinical trajectories in stroke survivors from European ancestry. These findings suggest that precision medicine approaches for stratifying stroke patients according to their genetic risk for elevated LDL-c have unique clinical utility in assessing post-stroke clinical management and should be further pursued.

## Acknowledgments

We gratefully thank all the participants in the VISP and UK Biobank studies without whom this research would not have been possible.

