## Supplementary figures and images for "Genetically-Determined Low-density Lipoprotein in Ischemic Stroke Survivors"

### Supplemental Figure

Flowchart for UKB:

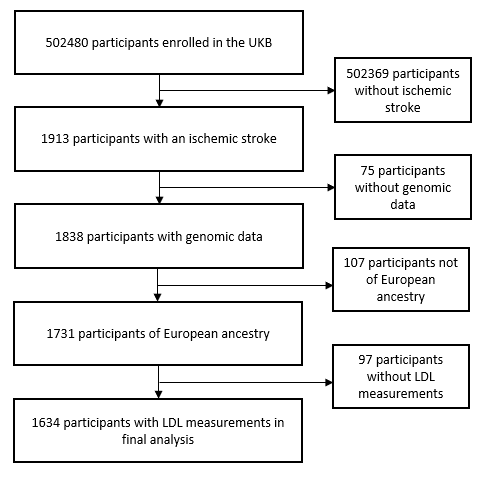
