## Supplemental Table 1 for "Genetically-Determined Low-density Lipoprotein in Ischemic Stroke Survivors"

Table 1. Baseline table for UKB:

| Variable | Polygenic susceptibility to high LDL-c | | | | |
| --- | --- | --- | --- | --- | --- |
|  | All | Low (0-20%) | Intermediate (20-80%) | High (80-100%) | P-value |
|  | n = 1634 | n = 327 | n = 981 | n = 326 |  |
| **Variables of interest:** | | | | | |
| LDL (mean, SD) | 105.87 (31.54) | 97.15 (28.35) | 107.34 (31.54) | 110.17 (33.06) | <0.001 |
| **Demographics:** | | | | | |
| Age at enrollment (mean, SD) | 60.78 (6.65) | 61.08 (6.47) | 60.69 (6.73) | 60.75 (6.59) | 0.653 |
| Female sex (n, %) | 518 (31.7) | 110 (33.6) | 326 (33.2) | 82 (25.2) | 0.018 |
| **Vascular risk factors:** | | | | | |
| Hypertension (n, %) | 1091 (66.8) | 218 (66.7) | 661 (67.4) | 212 (65.0) | 0.737 |
| Diabetes Mellitus (n, %) | 272 (16.6) | 74 (22.6) | 160 (16.3) | 38 (11.7) | 0.001 |
| Smoking (n, %) | | | | | |
| Never | 617 (37.9) | 120 (36.8) | 380 (38.9) | 117 (36.3) | 0.824 |
| Previous | 768 (47.2) | 153 (46.9) | 456 (46.6) | 159 (49.4) |  |
| Current | 241 (14.8) | 53 (16.3) | 142 (14.5) | 46 (14.3) |  |
| BMI (mean, SD) | 28.96 (5.01) | 29.39 (5.69) | 28.85 (4.92) | 28.89 (4.53) | 0.239 |
| **Comorbidities:** | | | | | |
| Prior MI (n, %) | 200 (12.2) | 22 (6.7) | 127 (12.9) | 51 (15.6) | 0.001 |
| CHF (n, %) | 83 (5.1) | 16 (4.9) | 48 (4.9) | 19 (5.8) | 0.789 |
| **Medications:** | | | | | |
| Statin use (n, %) | 1225 (75.0) | 236 (72.2) | 743 (75.7) | 246 (75.5) | 0.424 |
