## Supplemental Table 2 for "Genetically-Determined Low-density Lipoprotein in Ischemic Stroke Survivors"

Table 2. Unadjusted analysis

| **Uncontrolled and Resistant Hyperlipidemia: unadjusted** | | | | |
| --- | --- | --- | --- | --- |
| Garnet-VISP | | | | |
| Genetic Risk Cohort (n) | Uncontrolled | | Resistant | |
|  | n (%) | *p* | n (%) | *p* |
| Low (313) | 198 (63.3) | 0.003 | 54 (17.3) | 0.035 |
| Intermediate (940) | 680 (72.3) |  | 225 (23.9) |  |
| High (314) | 232 (74.0) |  | 76 (24.2) |  |
| Total (1567) | 1110 (70.8) |  | 355 (22.7) |  |
| UKB | | | | |
| Genetic Risk Cohort (n) | Uncontrolled | | Resistant | |
|  | n (%) | *p* | n (%) | *p* |
| Low (327) | 132 (40.4) | <0.001 | 64 (19.6) | <0.001 |
| Intermediate (981) | 528 (53.8) |  | 348 (35.5) |  |
| High (326) | 187 (57.4) |  | 125 (38.3) |  |
| Total (1634) | 847 (51.8) |  | 537 (32.9) |  |
